# Do Lockdowns Bring about Additional Mortality Benefits or Costs? Evidence based on Death Records from 300 Million Chinese People

**DOI:** 10.1101/2020.08.28.20183699

**Authors:** Jinlei Qi, Dandan Zhang, Xiang Zhang, Peng Yin, Jiangmei Liu, Yuhang Pan, Tanakao Takana, Peiyu Xie, Zhaoguang Wang, Shuocen Liu, George F. Gao, Guojun He, Maigeng Zhou

## Abstract

**Objectives:** To estimate the short-term effect of stringent lockdown policies on non-COVID-19 mortality and explore the heterogeneous impacts of lockdowns in China after the COVID-19 outbreak.

**Design:** Employing a difference-in-differences method.

**Setting:** Using comprehensive death records covering around 300 million Chinese people, we estimate the impacts of city and community lockdowns on non-COVID-19 mortality outside of Wuhan.

**Participants:** 44,548 deaths recorded in 602 counties or districts by the Disease Surveillance Point System of the Chinese Center for Disease Control and Prevention from 1 January 2020 to14 March 2020.

**Results:** We find that lockdowns reduced the number of non-COVID-19 deaths by 4.9% (cardiovascular deaths by 6.2%, injuries by 9.2%, and non-COVID-19 pneumonia deaths by 14.3%). A back-of-the-envelope calculation shows that more than 32,000 lives could have been saved from non-COVID-19 diseases/causes during the 40 days of the lockdown on which we focus.

**Main outcome measures:** Weekly numbers of deaths from all causes without COVID-19, cardiovascular diseases, injuries, pneumonia, neoplasms, chronic respiratory diseases, and other causes were used to estimate the associations between lockdown policies and mortality.

**Conclusions:** The results suggest that the rapid and strict virus countermeasures not only effectively controlled the spread of COVID-19 but also brought about unintended short-term public health benefits. The health benefits are likely driven by significant reductions in air pollution, traffic, and human interactions during the lockdown period. These findings can help better inform policymakers around the world about the benefits and costs of lockdowns policies in dealing with the COVID-19 pandemic.

## Introduction

By the end of 2020, COVID-19 had affected more than 219 countries and caused more than 1.7 million deaths worldwide.^1^ Facing this unprecedented crisis, different countries adopted various measures to mitigate its impacts, ranging from one extreme, where governments imposed draconian measures to restrict human mobility immediately after the outbreak, to the other extreme, where governments were reluctant to adopt any serious disease preventive measures and explicitly resorted to herd immunity. Effective policies not only depend on the social preferences of people and the capacity of government but also depend on our accurate understanding of the costs and benefits of different counter-COVID-19 measures. However, relatively little is known about the broader impacts of these policies.

A key component when evaluating the welfare implications of the anti-contagion policies is their overall public health consequences. Multiple studies have shown that strict social distancing and human mobility restrictions can effectively control the spread of COVID-19 and thus save lives from the virus.^2-6^ However, it remains unknown to researchers and policymakers how such interventions affect disease patterns and deaths from other causes. On the one hand, hospitals may decline non-urgent service requests (especially when the system is overburdened by COVID-19)^7^ and the fear of getting infected by COVID-19 may make patients reluctant to visit hospitals. This could impact the quality of health services and delay medical treatment, which would negatively affect population health. Additionally, in many countries, the strict virus containment policies led to sudden and sharp economic disruption, causing massive layoffs.^8^ As documented in the previous literature, such economic downturns and high unemployment could also damage population health.^9-15^ These factors would increase mortality when strict counter-virus measures were enforced. On the other hand, because the virus containment policies significantly improved air quality, restricted human-to-human interactions, and reduced work and traffic accidents, it is also possible that a large number of people could be saved from dying from air pollution, other types of communicable diseases, and accidents.^17-19^ Therefore, it is of great scientific and policy relevance to assess whether the counter-virus measures bring about additional public health gains or additional public health losses.

Using data from China, we examine how city and community lockdown policies affect non-COVID-19 mortality. We focus on China because the country mandated strict social distancing and lockdown policies to control the virus. Within a few weeks after the COVID-19 outbreak in Wuhan, a large number of cities enforced strict quarantines, traced close contacts, prohibited public gatherings, mandated social distancing, and limited human mobility. A large number of cities were locked down even though they had less than 100 confirmed cases (Figure SM1 and Figure SM2). Exploiting the staggered introduction of city and community lockdowns in different cities of China, we estimate the impacts of lockdowns on the number of deaths from various causes and explore the channels through which these impacts are manifested. These results will help policymakers around the world design effective measures to mitigate the damages from the pandemic.

The core of our empirical analysis uses the comprehensive deaths record from China’s Disease Surveillance Points (DSPs) system, covering more than 324 million people in 605 DSP’s districts/counties in 321 cities, which accounts for 24.3% of the country’s population.^20, 21^ This dataset includes cause-specific deaths, which allows us to examine the mechanism of lockdowns’ impacts on non-COVID-19 mortality. Each city’s lockdown information is collected from news media and government announcements. During the end of January and the mid of February, a large number of Chinese cities have implemented the lockdown policies (Figure 1). There are two types of lockdowns: city lockdown and community lockdown. The former is defined as mobility being restricted across different cities, and the latter is defined as the restriction of mobility within a city. Matching these datasets, we construct a daily DSP site-level panel dataset from January 1 to March 14, 2020, which is the period largely overlapping with the coronavirus outbreak in China. Our dataset includes 393,133 death records that were reported to the DSPs system by May 15, 2020 (Table SM1). Note that we exclude 3 DSPs in Wuhan from the baseline analysis because the city is the epicenter of the outbreak in China, and we are concerned that its death reporting process could have been affected during the study period.^22^

To quantify the impacts of lockdowns on mortality, we employ a difference-in-differences (DiD) approach, which is an econometric approach and is widely used to infer causal impacts of various policies and events using observational data^23^. An advantage of this approach is that it compares the policy effects relative to the plausible counterfactuals. While the results from a before-and-after comparison could be driven by different mortality trends or other unobserved confounders, DiD compares the changes in mortality between the locked-down DSPs (treatment group) and the non-locked-down DSPs (control group) before and after the enforcement of lockdown policies. In other words, the control group can serve as a counterfactual, mimicking what would have happened in locked down DSPs in the absence of the lockdown, which essentially allows us to compare the policy effects relative to business as usual. Note that a key assumption of the DiD is that the treatment and the control group follow parallel trends in the number of deaths in the absence of the lockdown policies. We examine whether this assumption is likely to hold using an event-study test. We describe the model in more detail in the Materials and Methods.

## Methods

### Materials

#### Study area

We collected 44,548 deaths from 605 Disease Surveillance Point (DSPs) districts/counties from January 1 to March 14, 2020, which were reported to the DSPs system by May 15, 2020. In our baseline analysis, we exclude three points (districts) in Wuhan due to concerns that the data might be unrepresentative because the pandemic started there.

#### Mortality Data

Weekly mortality data are provided (See supplementary materials and methods). The causes of death are coded in accordance with the International Classification of Diseases-10th revision (ICD-10). We classified the main underlying causes of deaths into 6 categories: I00-I99 for cardiovascular diseases (CVD), V01-Y89 for injuries, J12-J15, J18.9 and J98.4 for pneumonia (excluding COVID-19), C00-C97 for neoplasms, J30-J98 for chronic respiratory diseases, and other causes (remaining ICD-10 codes for all other causes). We further disaggregate cardiovascular diseases, injuries, and pneumonia deaths into specific diseases/causes. Cardiovascular diseases include stroke (I60-I62, I67, and I69), myocardial infarction (I20-I25), and other cardiovascular diseases. Injuries include traffic accidents (V01-V04, V06, V09, V87, V89, and V99), suicide (X60-X84 and Y87), and other injuries. Pneumonia includes mycoplasma pneumonia (J18.9), viral and bacterial pneumonia (J12-J15), and pulmonary infection (J98.4). We also divide the daily number of deaths into three age groups (0-15, 15-64, and ≥65). All death data are analyzed at the aggregated level.

#### Lockdown Data

We collected local governments’ lockdown information city by city from news media and government announcements, details in supplementary materials and methods. The evolution of different DSPs’ lockdown status is presented in Figure SM1 and Figure SM2. In Table SM11, we further provide a complete list of cities that adopted different lockdown policies at different times. The lockdowns gradually spread to different surveillance districts/counties between January 23 and February 20. By the end of February, 486 out of 602 surveillance points had lockdown policies.

#### Weather Data

Weather variables include daily temperature, atmospheric pressure, relative humidity, wind speed, and precipitation. The data are obtained from the China Meteorology Administration (CMA). We aggregate station-level air pollution data to city-level data using the inverse squared distance (to city centers) as the weights. Stations closer to the population center are given higher weights so that city-level weather data can be representative of people dwelling in the city.

#### Air Pollution Data

We obtain air pollution data from the Ministry of Ecology and Environment. The original dataset includes hourly air quality readings from over 2,000 monitoring stations covering 338 prefectural cities in China. We follow the same procedure to aggregate station level air pollution data to the city level. As an omnibus measure of the overall air quality, we use PM_2.5_ concentration in our regressions. Our results are quantitatively unchanged if we use the Air Quality Index or PM_10_.

#### Socio-Economic Conditions

We assemble the socio-economic data at the city or county level from the 2018 China City Statistical Yearbook and 2018 China County Statistical Yearbook, including GDP, population, and the number of hospital beds per 1,000 people. We also obtain data on the employment share of the manufacturing and service industries using the 10% sample of the 2015 1% Population Sampling Survey in China.

#### Summary Statistics

We report the summary statistics of mortality, lockdown status, and other covariates for 602 DSP counties in Table SM1. In Panel A, we report the summary statistics of the DSPs data. The average daily total number of deaths at the county level is 8.7, with a standard deviation of 0.025. The leading cause of death during this period is cardiovascular diseases, which account for 49.7% of all deaths. The second leading cause of death is neoplasms (22.3%), followed by chronic respiratory diseases (8.7%), and injuries (5.5%). In Panel B, we report the summary statistics of several other variables. The PM_2.5_ concentration during our study period is 50 µg/m^3^, five times higher than the WHO standard (10 µg/m^3^ for annual mean, and 25 µg/m^3^ for a daily mean). The average share of employment in the manufacturing industries was 24.2% as of 2015.

### Statistical Analysis

To quantify the impacts of lockdowns on mortality, we employ a difference-in-differences (DiD) approach, which is an econometric approach and is widely used to infer causal impacts of various policies and events using observational data^23^. An advantage of this approach is that it compares the policy effects relative to the plausible counterfactuals. While the results from a before-and-after comparison could be driven by different mortality trends or other unobserved confounders, DiD compares the changes in mortality between the locked-down DSPs (treatment group) and the non-locked-down DSPs (control group) before and after the enforcement of lockdown policies. In other words, the control group can serve as a counterfactual, mimicking what would have happened in locked down DSPs in the absence of the lockdown, which essentially allows us to compare the policy effects relative to business as usual. Note that a key assumption of the DiD is that the treatment and the control group follow parallel trends in the number of deaths in the absence of the lockdown policies. We examine whether this assumption is likely to hold using an event-study test. We describe the model in more detail in the Supplementary Materials and Methods.

### Ethical Approval

The ethics committee from the National Center for Chronic Non-Communicable Disease Control and Prevention (NCNCD) of the Chinese Center for Disease Control and Prevention approved the study. No individual consent was required as all the data were analyzed at aggregated level, and no patients were involved in setting the research question or the outcome measures, nor were they involved in developing plans for recruitment, design, or implementation of the study.

## Results

### Impacts of City and Community Lockdowns on Non-COVID-19 Deaths

Figure 2 summarizes the baseline regression results by fitting the DiD model (Equation A1; full results are in Table SM2). Panel A reports the effects on the number of deaths, while Panel B reports the percentage change. In row (1), we find that lockdowns overall have a negative impact on non-COVID-19 mortality. After human mobility is restricted, the DSP-level daily number of deaths decreased by 0.429 (or 4.92%), as compared to the control group.

In rows (2) to (7), motivated by several factors that could potentially affect population health during the lockdown period, we separately examine the effects on different causes of death. We are especially interested in the following three outcome variables: cardiovascular diseases (CVD), injuries, and (non-COVID-19) pneumonia deaths. Existing literature on the acute effects of air pollution suggests that elevated air pollution levels can significantly increase deaths from strokes, myocardial infarction, and other types of cardiovascular diseases.^24-25^ We thus expect the number of deaths from CVDs may decrease due to the improved air quality.^27^ As shown in row (2) of Figure 2, we find that cardiovascular deaths were reduced by 6.2% (0.27 in levels) after lockdown. Relatedly, as the lockdown policies restrict production, social activities, and traffic, we expect the number of deaths from injuries (which include workplace injuries, traffic accidents, etc.) to also drop. The result in row (3) of Figure 2 confirms this conjecture; we observe that the number of deaths caused by injuries decreased by 9.2% (0.044 in levels). In addition, as human mobility is greatly restricted during the lockdown period, this should reduce the likelihood of people getting infected by and dying from other types of bacteria and viruses that cause pneumonia. The result in column (4) shows that deaths from non-COVID pneumonia were reduced by a large margin of 14.7% (0.022 in levels) during the lockdown period.

In rows (5) to (7), we report the findings on several other causes of death that are less likely to be affected by short-term restrictions on human activities. They include deaths from neoplasms, chronic respiratory diseases, and other diseases. While the coefficients for these causes of death are also negative, they are all not statistically significant. We thus conclude the temporary human mobility restrictions during China’s lockdowns primarily reduce the deaths caused by acute diseases and accidents and have a weaker impact on people with chronic diseases and cancers.

Some additional analyses complement our main findings. A key assumption of the DiD is that the treatment and the control group follow parallel trends in the number of deaths in the absence of the lockdown policies. Using an event-study approach, we show that this assumption is likely to be held (Figure 3 and Supplementary Note 1 and Table SM3). Also, we find that our results are robust to the inclusion of additional controls, adoption of different weighting, and sampling (Supplementary Note 2, Table SM5, and Figure SM3). Finally, we further disaggregate the data into more specific causes/diseases (Table SM4). For example, in the cardiovascular disease category, we observe that deaths from myocardial infarction, strokes, and other types of cardiovascular diseases all significantly decreased after the lockdown.

### Heterogeneity

In Figure 4, we examine the heterogeneous impacts of lockdowns on mortality. Here we report our findings on the total number of non-COVID-19 deaths and explore the following dimensions: baseline income (measured by per capita GDP in 2018), healthcare resources (measured by hospital beds per thousand people in 2018), air pollution levels (measured by average PM_2.5_ concentrations in 2019), industrial structure (measured by the share of employment in manufacturing industries in 2015), and initial health status (measured by mortality rate in 2019).

To do so, we interact the lockdown indicator separately with each of the heterogeneity dimensions in the regression (Table SM6), and then plot the predicted impacts and their 95% confidence intervals in Figure 4. We observe significant heterogeneities with respect to the air pollution level, the employment shares in the manufacturing industries, and the baseline mortality level. Specifically, the health benefit of lockdowns on mortality is greater when a DSP is more polluted and more industrialized, and when the initial health status is worse. This finding is consistent with several previous studies which show that China’s lockdowns significantly reduced the air pollution.^17-18^

We also repeat this exercise separately for deaths from specific causes: cardiovascular diseases, injuries, and non-COVID-19 pneumonia (Figure SM4). Several patterns stand out: (1) for cardiovascular diseases, there exist significant heterogeneities for air pollution and industrial structure, with more polluted and more industrialized cities seeing fewer deaths from cardiovascular diseases during lockdowns relative to other cities (Panel a); (2) for injuries, the more industrialized the DSP, the higher its initial injury mortality, and, as expected, the greater the impact of the lockdown (Panel b); (3) for pneumonia, we only observe significant heterogeneity with respect to initial mortality rate, *i*.*e*., cities with a higher initial pneumonia mortality rate are more strongly affected by lockdowns (Panel c). Across all the causes of death, per capita GDP and availability of healthcare resources do not seem to play an important role in terms of magnitude, although occasionally they are statistically significant. The corresponding regression results are reported in Tables SM7-9. As a side note, we also examined many other dimensions of heterogeneity, including the severity of the COVID-19 outbreak, alternative measures of health care resources, other measures of economic structure, etc. However, we do not observe strong heterogeneities along these dimensions and thus do not report them in the paper.

Finally, we investigate which age group(s) are driving the overall reduction in mortality. We expect older people and younger people to be sensitive to the overall lockdown policies, while we expect adults to be vulnerable to injuries and accidents. Figure SM5 summarizes the results. We find that children (−10.6% in row 1) and the elderly (−5.5% in row 5) are indeed more likely than adults (−2.5% in row 2) to be saved by the lockdown policies. If we further examine different causes of death, we find that the elderly is saved both from air pollution-related disease (−6.6% in row 6) and infectious disease (−17.0% in row 8), and younger adults are protected from injuries (−14.7% in row 4). These results are generally consistent with our understanding of the threats of various diseases to different age groups. More detailed results are represented in Table SM10.

### Back-of-the-envelope calculation

In Figure 5, using the estimates in our analyses, we calculate the averted non-COVID-19 deaths in the whole nation due to the lockdown policies during our study period. In Panel a, we plot the predicted average daily deaths. The red and blue lines respectively represent the predicted deaths with and without lockdown policies. Therefore, the differences between these lines can be regarded as the lockdown effects. We see that these two lines start to diverge as more cities implement lockdown policies, and the difference remains stable throughout mid-March.

Because our dataset includes around a quarter of the Chinese population, we apply our estimates to the entire Chinese population in Panel b. During our study period, 486 DSPs (80.7%) eventually implemented lockdowns, with an average of 38.5 days. We apply our estimates to all the cities that implemented the lockdown policies and calculate the number of averted deaths during our study period. We find that the lockdown policies brought about considerable health benefits: as many as 32,023 lives may have been saved. If we look at the cause-specific effects, we find that cardiovascular diseases account for 62.9% (20,129) of overall averted deaths. Deaths from injury also declined by 10.2% (3,261), pneumonia by 5.0% (1,607), respiratory by 7.4% (2,373), and cancer by 8.5% (2,726).

## Discussion

When COVID-19 spread across the globe, we observed a large variation in the public responses in mitigating its impacts: some countries immediately adopted harsh counter-virus measures while others delayed the launch of the policies. As an example of prompt and stringent responses to the COVID-19 outbreak, we investigate the mortality consequences of community and city lockdowns using data from China (excluding Wuhan) during the pandemic period. Here, we discuss several important implications of our findings.

First and foremost, our findings demonstrate that the China’s lockdowns not only effectively controlled the spread of COVID-19, but also brought about unintended short-term benefits to population health during this period. We find that such policies reduced non-COVID-19 deaths by 4.92%, which corresponds to 32,000 averted deaths in the nation during 40 days of lockdown. Given the increasingly heated cost-benefit debates regarding different counter-COVID-19 policy choices across the world, our results provide a benchmark to understand the health consequences of the lockdown policies. Besides China, several other countries have managed to take the COVID-19 threat under control after one to two months’ strict social distancing, largely because they dealt with the COVID-19 quickly and decisively.

Second, our research points out the directions to improve population health after the pandemic. In particular, we observe a significant reduction in the number of cardiovascular deaths during the lockdown periods, and the effect is larger in cities with higher levels of initial air pollution. A back-of-the-envelope calculation suggests that the total number of averted premature deaths from cardiovascular diseases in the locked-down DSPs alone has far exceeded the total number of deaths caused by COVID-19 in China. This result suggests that air pollution imposed a significant health risk to the Chinese population and it is critically important for the government to continue to improve the environmental quality even when the lockdown is lifted.^28, 29^ Besides, the finding on pneumonia mortality confirms that reducing human contacts and raising awareness of preventive measures (such as wearing masks) not only helps control the spread of COVID-19, but also other infectious diseases. These measures should be more appreciated by both public health practitioners and governments.

Third, our results also serve as corroborating evidence that China’s COVID-19 data outside of Wuhan are largely reliable. The logic is the following: if the deaths from COVID-19 were intentionally classified as other causes, such as pneumonia or other unclassified diseases, we might observe an unexplainable hike in those causes of death in the locked-down cities (presumably, there were few cases of COVID-19 in the control group). Our results suggest this is not the case; we find that the lockdown reduces all these causes of death in the locked-down cities (using data outside Wuhan), suggesting that COVID-19 deaths are unlikely to be misreported in a substantial way. For Wuhan, however, we do have suggestive evidence of potential misclassification of COVID-19 deaths, as including Wuhan in the regression reverses the sign for deaths from non-COVID-19 pneumonia.

Finally, while the literature has emphasized that economic downturns are usually associated with increased mortality (particularly in less affluent countries), our analyses show that the negative health effects of income shocks during China’s lockdowns were offset by unintended benefits to population health, at least in the short run. While economic collapse is likely to seriously harm public health in the long run, we believe that countries currently affected by COVID-19 can maintain overall population health for a short time by containing the virus as quickly as possible through strict social distancing/mobility restrictions.

### Strengthen and limitations of study

This study has several major strengths. First, we use the largest mortality database in China, covering a quarter of the Chinese population. The data are nationally representative, and our findings are unlikely to be affected by potential data misreporting in a specific city (i.e., Wuhan). Second, this study examines the effects of lockdowns on a variety of causes of death in China, which provides insights on the overall health implications of lockdown policies. Third, we explore the mechanism of lockdowns’ impacts on cause-specific deaths and show that improvement in air quality, reduction in accidents, and less human interactions can help explain the impacts.

However, a few limitations should be noted here. First, due to the data unavailability in personal daily activities and family care at the household level, we are unable to study the effect of lockdowns on morbidity, especially for those who were concerned and did not go to hospitals when getting sick. Second, our research only focuses on the short-term effect of the lockdown policies, so our findings cannot be applied to the long-term case. In the long run, there could be a harvesting effect (normally negative) on mortality rates, especially for mortality due to chronic diseases, which might be caused by the delay of medication and physical examinations during lockdowns. Also, if lockdowns were sustained for an extended period of time, low-income households would suffer a lot, and their health conditions would significantly deteriorate. Finally, we focus on China, where households generally have high saving rates and support stringent virus-control policies. Our findings are thus more relevant to countries with similar institutions, such as Japan and South Korea. Future research using data from other countries is needed to better understand the overall benefits and costs of the global pandemic.

## Conclusions and implications

Understanding the broader social and health impacts of different counter-COVID-19 policies is critical for optimal policy design. Using comprehensive death records data from China, this paper provides the first empirical evidence that strict city and community lockdowns brought about unintended short-term health benefits. We observe fewer deaths from cardiovascular diseases, traffic accidents, and non-COVID-19 pneumonia during the lockdowns. This result is likely to be driven by the significant improvement in air quality, reduction in traffic volume, and less human interactions. Policymakers in other countries, particularly those face similar public health challenges, should consider these unintended benefits in designing their strategies to fight against COVID-19.

## Data Availability

The DSPs data are proprietary data owned by the Chinese CDC. They can be accessed through application to the National Center for Chronic and Noncommunicable Disease Control and Prevention (a subsidiary of the Chinese CDC). The codes necessary to re-produce all the tables in the paper are ready to submit to the journal as supplementary materials or to post on a public repository.

## Contributors

M.Z., G.H. and G.G. are joint senior authors. M.Z., G.H., D.Z. and J.Q. designed the project. J.Q., D.Z., X.Z., P.X., Z.W., and S.L. curated data. D.Z., J.Q., X.Z., P.Y., J.L, and T. T., analyzed the data. M.Z., G.H, D.Z. and J.Q. interpreted the results. G.H., D.Z., J.Q, and T.T. wrote the manuscript. G.H., G.G., J.Q., D.Z., and P.Y. edited the manuscript.

## Acknowledgments

We thank all the staff who work in the primary health facilities, hospitals, and Center for Disease Control and Prevention for death reporting at county/district, city, province, and national levels. We also thank Yun Qiu for sharing the community lockdown data with us. Wei Wang and Yaxuan Liu provided for research assistance.

## Supplementary Materials and Methods

### Materials

#### DSP Data

The Disease Surveillance Points (DSPs) system is managed by the Chinese Center for Disease Control and Prevention. The system collects death records from the surveillance locations to understand death and disease patterns in China. The system was established in 1978 and has gradually increased its geographical coverage over the past four decades. In 2013, the system got a major upgrade and expanded its coverage from 161 points to 605 points, making the data representative both at the provincial and national levels. Each surveillance point represents a district (if in urban areas) or a county (if in rural areas). For each surveillance point, deaths that occurred in both hospitals and homes are reported, and the causes of death are determined according to a standard protocol by trained staff located in local hospitals or CDC branches. The DSPs system covers more than 324 million people in China, which accounts for 24.3% of the country’s population.^20,21^ The quality control procedures include annual training of standard workflow, random checking of the accuracy of disease classification and duplication, retrospective surveys on underreporting, and logic checks on the completeness and accuracy of disease codes. These quality checks are required to be done at the county, province, and national levels.

In addition to the DSPs system, the Chinese CDC also manages the Communicable Disease Surveillance (CDS) system. The DSPs system is used for death registration, while the CDS system is used to monitor the development of infectious diseases, including COVID-19. These two systems are managed by different subsidiaries in the Chinese CDC and have separate administrative structures. Because the purpose of the DSPs system is to understand the overall cause-of-death patterns in China, and because the death registration process carries no political stake, there is no incentive for local hospitals or CDC branches to hide regular death information from the central government.

#### Details of Lockdown Data

Most of the cities’ lockdown policies were directly issued by the city-level governments, while a few were promulgated by the provincial governments. There are two types of lockdowns: city lockdown and community lockdown. The former is defined as human mobility being restricted across different cities, and the latter is defined as mobility being restricted within a city. At the early stage of the outbreak, to prevent the virus from spreading outside Hubei province, city lockdowns were adopted in Wuhan and its neighboring cities. The purpose of city lockdowns was to restrict people in the epicenter of coronavirus from traveling to other cities. Later, as more cases were identified in other cities, community lockdowns were implemented to further control the spread of the coronavirus within cities. The time lag between city lockdowns and community lockdowns was typically one to two weeks.

### Methods

We use a generalized Difference-in-Differences (DiD) model to identify the impact of counter-COVID-19 measures on mortality. First, in our baseline regression, we estimate the relative change in the number of deaths between the treated and control DSPs using the following model:

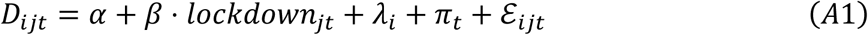

where *D*_*ijt*_ denotes the daily number of deaths in DSP *i* in city *j* on date *t*, and *lockdown*_*jt*_ is a dummy variable indicating whether a city/community lockdown is in place in city *j* on date *t*. The lockdown dummy takes the value one if either city lockdown or community lockdown was implemented, and zero otherwise. Thus, the coefficient *β* measures the average effect of three types of lockdown policies: mobility restrictions across cities (city lockdown), mobility restrictions within a city (community lockdown), and both restrictions (city lockdown + community lockdown). To understand how the city and community lockdowns affect health outcomes differently, we separately estimate these effects (Table SM5). *λ*_*i*_ are DSP-fixed effects and *π*_*t*_ indicate date fixed effects. *ε*_*ijt*_ is the error term.

The county fixed effects, *λ*_*i*_, which are a set of DSP-specific dummy variables, can control for time-invariant confounders specific to each DSP. For example, the DSP’s geographical conditions, short-term industrial and economic structure, income, and natural endowment can be controlled by introducing the DSP fixed effects. The date fixed effects, *π*_*t*_, are a set of dummy variables that account for shocks that are common to all DSPs in a given day, such as the nationwide holiday policies, macroeconomic conditions, and the national time trend for mortality. As both location and time fixed effects are included in the regression, the coefficient *β* estimates the difference in the number of deaths between the treated (locked down) and the control cities before and after the enforcement of the lockdown policy. We also add a set of control variables in the regressions to check the robustness of the results (Figure SM3).

The underlying assumption for the DiD estimator is that lockdown and control cities would have parallel trends in the number of deaths in the absence of the event. Even if the results show that mortality declines in the treatment counties after the lockdown, the results may not be driven by the lockdown policy, but by systematic differences in treatment and control cities. This assumption is untestable because we cannot observe the counterfactual: what would happen to the mortality levels in the locked-down counties if such policies were not enforced. Nevertheless, we can still examine the trends in mortality for both groups before the lockdown and investigate whether the two groups are indeed comparable. To do so, we conduct the event study and fit the following equation:

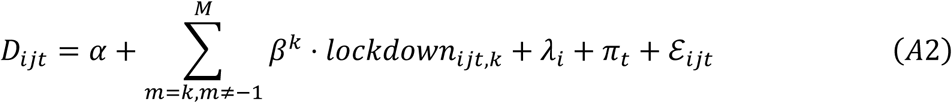

where *lockdown*_*jt,k*_ are a set of dummy variables indicating the treatment status at different periods. Here, we put 7 days (one week) into one bin (*bin m ∈ M*), so that the trend test is not affected by the high volatility of the daily number of deaths.

The dummy for *m* = −1 is omitted in Equation (A2) so that the post-lockdown effects are relative to the period one week before the launch of the policy. The parameter of interest *β*^*k*^ estimates the effect of lockdown *m* weeks after the implementation. We include leads of the treatment dummy in the equation, testing whether the treatment affects the air pollution levels before the launch of the policy. Intuitively, the coefficient *β*^*k*^ measures the difference in the number of deaths between cities under lockdown and otherwise in period *k* relative to the difference two weeks before the lockdown. If lockdown reduces mortality, *β*^*k*^ would be negative when *k* ≥ −1. If the pre-treatment trends are parallel, *β*^*k*^ would be close to zero when *k* ≤ −2.

We feel confident in using the estimates from our main results to calculate the averted deaths in the entire country, because our dataset includes around one-quarter of the Chinese population and are representative. To do so, we predict the number of deaths in two scenarios: with/without lockdown policies. Taking the difference between these two predicted deaths, we can calculate the number of saved lives from the lockdown policies. To do so, we first predict the number of deaths with lockdown policies in each DSP county/district in each day by fitting the following model:

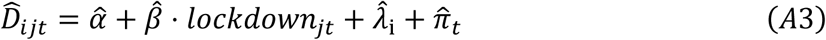

where 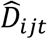 denotes the predicted deaths with lockdown policies in each DSP county/district *i* in city *j*.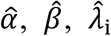 and 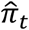 are the fitted values from Equation (A1). In this function, predicted deaths in each DSP, denoted by 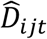, can be affected by the lockdown status (represented by *lockdown*_*jt*_).

We then predict the counterfactual, i.e., the number of deaths that would have occurred without lockdowns in any DSP, by fitting the following equation:

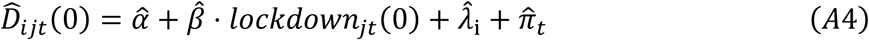

where 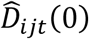 denotes the predicted averted deaths without any lockdown policies. *lockdown*_*jt*_ (0) always takes a value of zero so that this function is not affected by the policies. Taking the differences between 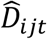 and 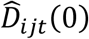, we can calculate how many non-COVID-19 deaths are saved from the lockdown policies in each DSP in each day.

Because lockdowns were implemented for 38.5 days on average, we estimate the following model to obtain the averted deaths in the whole country during our study period:

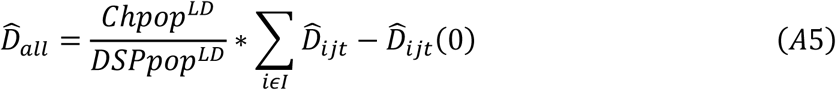

where 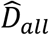 denotes the averted deaths in the entire county during our study period, *Chpop*^*LD*^ denotes the total Chinese population in locked-down cities (around 1,161 million), and *DSPpop*^*LD*^ represents the total population in locked-down DSPs counties/districts in our dataset (around 291 million in 486 DSPs). The difference between the scenarios with and without lockdowns, denoted by 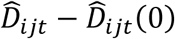, is totaled from January 1 to March 14, which is our study period (*i ϵ I*). Note that, in our main text, we repeat these steps to estimate the averted deaths from each cause and disease to understand how many averted deaths can be attributed to different diseases/causes.

## Supplementary Notes

**Supplementary Note 1: Tests for Pre-Treatment Parallel Trends**

The underlying assumption for the DiD estimates in Figure 2 is that lockdown and non-lockdown DSPs have parallel trends in mortality without the lockdown policies. To test how likely this assumption is to hold, we conduct an event study and investigate whether the two groups of cities have parallel pre-treatment mortality (Equation A2). Figure 3 plots our findings. Here we focus on the four outcomes (total deaths, and deaths from cardiovascular diseases, injuries, and pneumonia) that are statistically significant in Figure 2.

In Panel A, we compare the total number of non-COVID-19 deaths between the treatment and control groups before and after lockdowns. The difference between the two groups one week before the lockdown is set as the reference group (*i*.*e*., the zero coefficient for week-1), so the post-lockdown effects are relative to the period one-week before the launch of the policy. We do not observe systematic difference in the trends of mortality between the two groups one week before the city/community lockdown, *i*.*e*., the estimated coefficients for the lead terms (*k* ≤ −2) are positive or close to zero and statistically insignificant. This finding implies that the parallel trend assumption is likely to hold in our setting. In comparison, the trends break after the lockdown policies were enforced, *i*.*e*., the lagged terms (*k* ≥ 1) become negative and statistically significant. In addition, we observe that the difference becomes larger as more lags are included, suggesting an accumulating health benefit of city/community lockdowns.

In Panels B, D, and D of Figure 3, we repeat this exercise to investigate the trends in deaths from cardiovascular diseases, injuries, and non-COVID-19 pneumonia before and after lockdowns. The results suggest the parallel trend assumption holds for all these outcomes as well. The corresponding regression results are reported in Table SM3.

**Supplementary Note 2: Robustness Checks and Placebo Tests**

We conduct a variety of robustness checks and show that our results are not qualitatively affected by several decisions we make in the baseline analysis (Figure SM3 and Table SM5). First, we add weather variables into the baseline regressions, including daily average temperature, humidity, wind speed, and air pressure (R1), and find that the results are quantitatively unchanged. To further control for the differences in time trends between the treatment and control groups, we also include interactions of time-invariant variables (*i*.*e*., per capita GDP, number of hospital beds per thousand people, and total population) with a third-order polynomial function of time in the regressions (R2). The estimates remain similar. These results suggest that the lockdown policies are uncorrelated with these factors and lend additional credibility to our baseline findings.

Second, we weight regressions by population in each DSP (R3). Intuitively, this allows us to estimate the lockdown effects on an average individual, instead of an average DSP. Without weighting, cities with smaller populations could drive the baseline results. We find that the results remain very similar, suggesting that this is unlikely to be the case.

Third, we exclude 22 DSPs in Hubei province from the regression (R4). COVID-19 was first identified in Wuhan city in Hubei province. Thus, the DSPs in the province could be very different. We find that the results are quantitatively similar to the baseline. Next, we include the three DSPs (districts) located in Wuhan in the regressions and find the effects generally become weaker (R5). In addition, the sign for non-COVID-19 pneumonia is reversed and becomes positive (statistically insignificant) (Panel d). This finding suggests that some COVID-19 deaths could be misclassified as deaths caused by other types of pneumonia in Wuhan. Wuhan was the epicenter of COVID-19 in China and contributed to nearly half of the country’s COVID-19 cases. During the first few weeks after the outbreak, the city faced severe medical resource shortages, and many patients could not get immediate diagnoses and treatments. As a result, it would not be surprising to see that some people dying from COVID-19 in the city had been misclassified as dying from other types of pneumonia. In fact, a retrospective survey was recently conducted by the Chinese government, aiming to better classify causes of death in Wuhan. We thus exclude Wuhan from our baseline analysis.

Fourth, we separately estimate the effects of two types of lockdowns, *i*.*e*., city lockdowns and community lockdowns, on the number of deaths (Panel f in Table SM6). Shortly after the COVID-19 outbreak, a dozen cities around Wuhan (covering 65 DSPs) launched city lockdowns (mostly in late January). Later, as the virus started to spread outside Hubei province, more cities began to enforce community lockdown policies (including those that initially implemented city lockdowns). We include two policy dummies, *i*.*e*., city lockdowns and “city+community” lockdowns, into the regressions. The results show that “city+community” lockdowns play a more important role in reducing the number of total deaths, as well as deaths from cardiovascular diseases and non-COVID-19 pneumonia. In other words, restricting human mobility within cities seems critical to explaining our results. City lockdowns have an immediate effect on deaths from injuries, likely because people were not allowed to travel to other cities after city lockdowns.

Next, to address the concern that people die at home due to the lockdown might be uncounted and this miscalculation may overestimate the positive effect of the lockdown on public health, we repeat our analysis during the period of January 1 to March 14 based on the mortality data that was reported to DSPs system by April 23 and by June 15. If there were uncounted deaths due to the lockdown, there would be a larger number of death records for the later reported dates as more uncounted cases having been discovered. Thus, the lockdown effect will become weaker as the mortality data become more precise over time. However, the estimation results using the death records reported by different dates give very similar results, which implies that the mis-reporting problem in mortality is not a concern.

Finally, one potential threat to our empirical results is the large-scale travel across different cities during the study period. The COVID-19 outbreak coincided with China’s Spring Festival, during which many people leave the cities where they work and travel to their hometowns. If the death patterns are somehow correlated with this travel pattern, our results may be confounded. To address this issue, we conduct a placebo test using data from 2019 (Panel g in Table SM6). We assign the lockdown status to the same DSP in 2019 and compare changes in the number of deaths between the treatment and control DSPs before and after the “placebo” lockdowns. We find the “placebo” lockdowns do not have any impact on mortality in 2019, suggesting that our findings are not confounded by different mortality trends between the treatment and control groups related to the spring holiday travel.

## Supplementary Figures

**Figure SM1.**
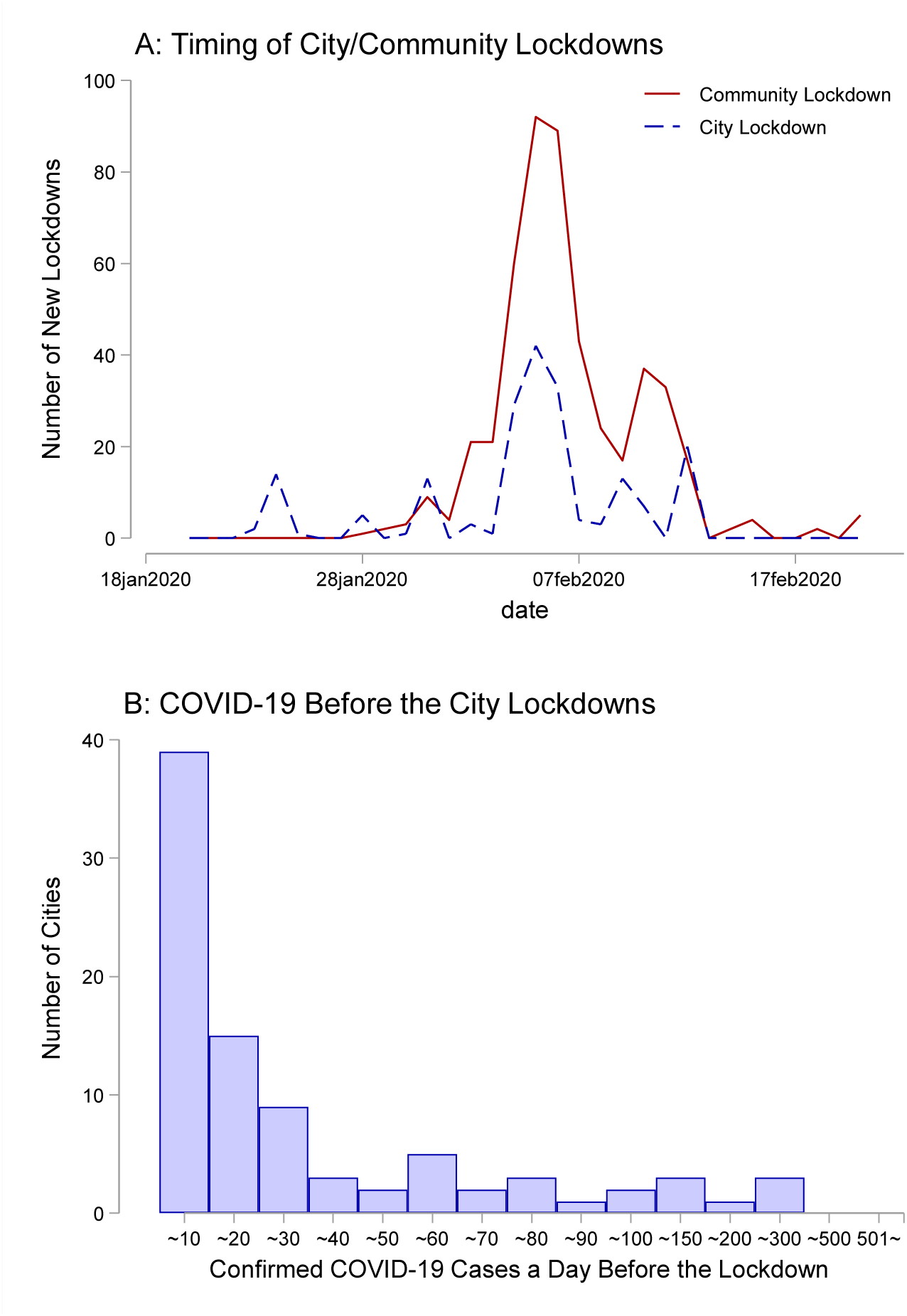
Distribution of lockdown dates for the DSPs. This graph shows the timing of the start of the city/community lockdowns other than Wuhan. Panel A reports the timing of community lockdowns and city lockdowns. In Panel B, we draw the distribution of confirmed cases a day before the implementation of city lockdowns. Because the confirmed COVID-19 cases are only available at the city level, the graph is based on city-level information.

**Figure SM2.**
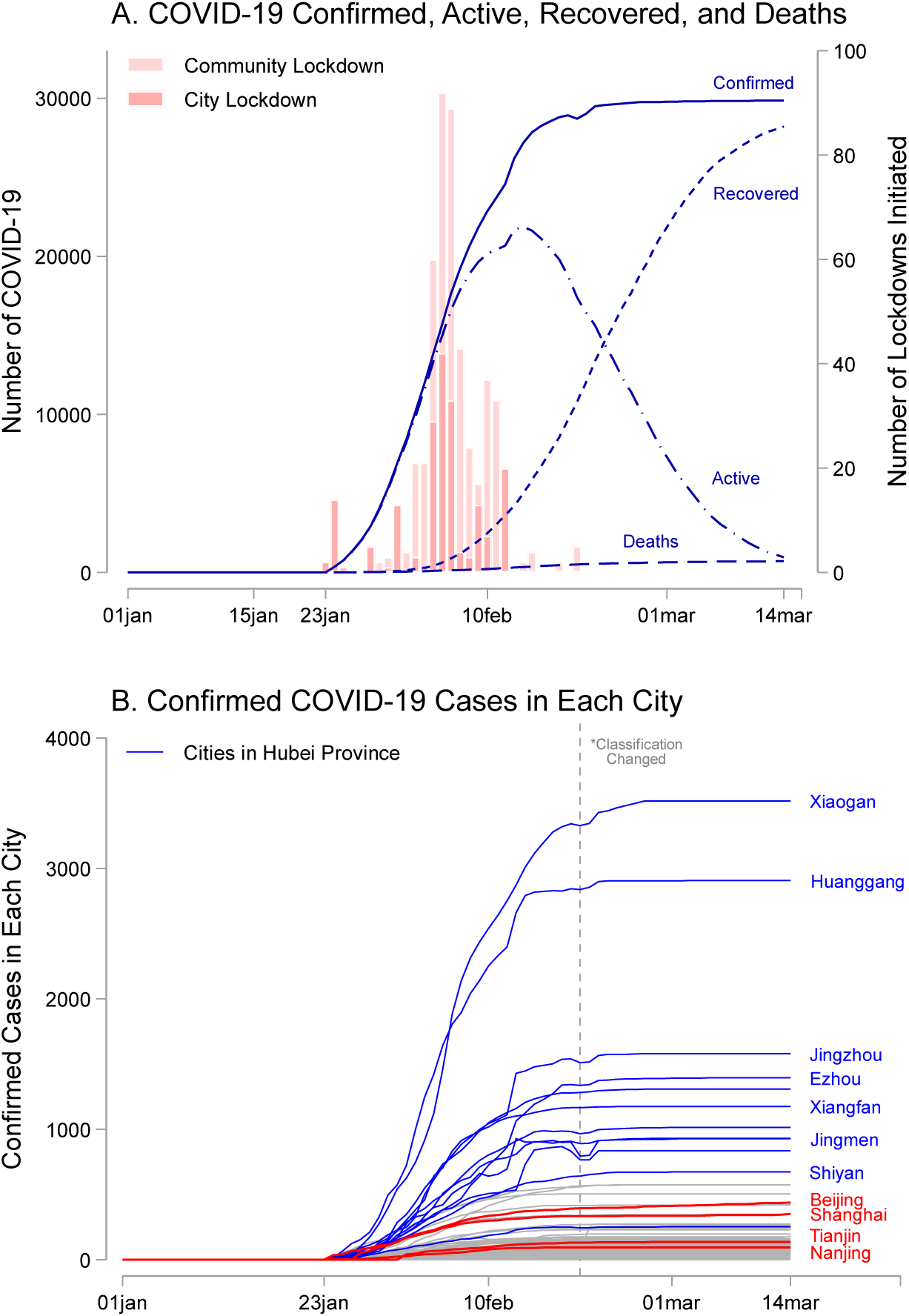
COVID-19 and city lockdowns outside of Wuhan in China. This graph shows the confirmed COVID-19 cases in each city. We drop data from Wuhan in both panels. Panel A describes the total confirmed cases, recovered cases, active cases, and deaths in the entire nation. The red bar graph shows the timing of the launch of lockdown policies. Panel B reports the confirmed cases in each city. The blue line denotes cities in Hubei province, and the red line denotes some other major cities in China. The grey dashed line indicates the date (February 20) when Hubei changed its diagnostic and reporting criteria on COVID-19.

**Figure SM3.**
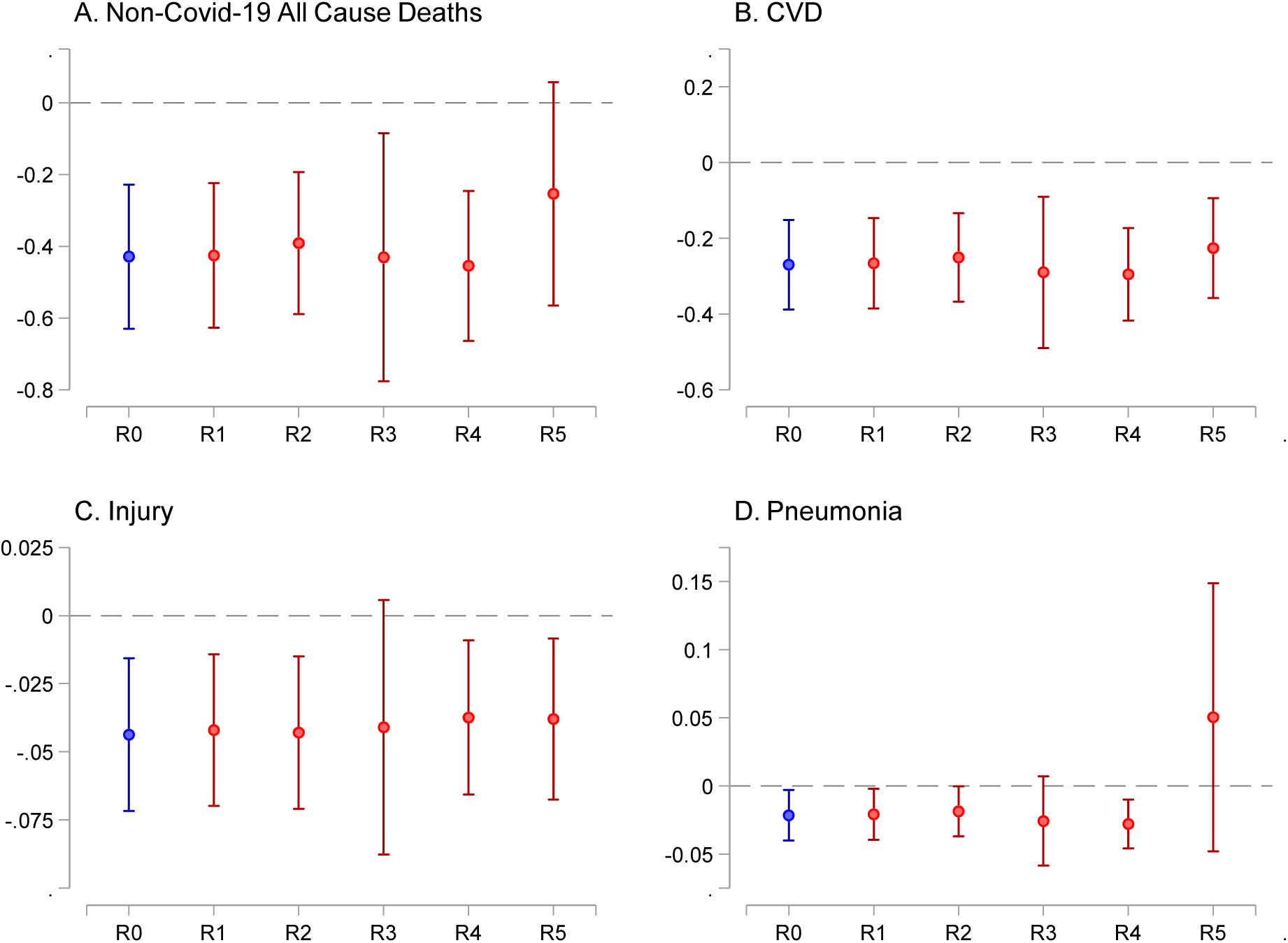
Robustness checks. Each column in each figure represents a separate DiD regression. In R0, we report the baseline results. In R1, the regression includes weather controls: daily average temperature, humidity, wind speed, and air pressure. R2 includes socio-economic status controls: interactions between time-invariant variables and a third-order polynomial function of time. In R3, the regression is weighted by the population. In R4, DSPs in Hubei province are excluded, while, in R5, three DSPs in Wuhan are included in the regression. DSP fixed effect and date fixed effect are both included in each regression. The standard errors are clustered at the DSP level.

**Figure SM4.**
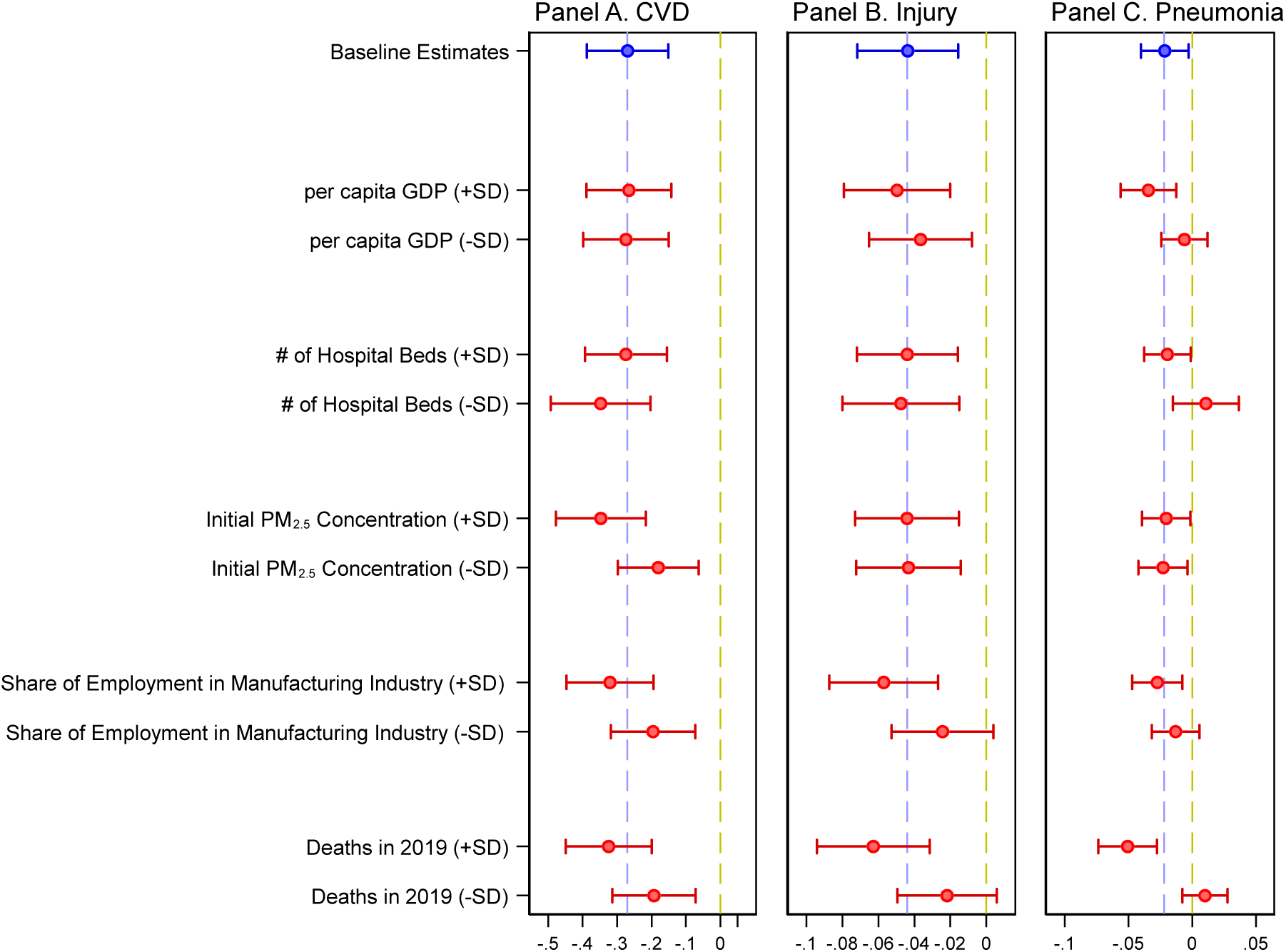
Heterogeneity analysis: CVD, injury, and pneumonia. Each row in the figure represent the predicted impacts of lockdown at different baseline socio-economic conditions, and their 95% confidence intervals. The heterogeneous dimension is shown in two scenarios: one standard deviation larger (+SD) / smaller (−SD) than mean. The prediction is based on the estimates from Table SM6-8. The top blue dot and line represent the baseline point estimates and 95% confidence interval for each disease/cause category, respectively. DSP fixed effect and date fixed effect are both included in each regression. The total number of observations for each regression is 44,548 covering 602 DSP counties. The standard errors are clustered at the DSP level.

**Figure SM5.**
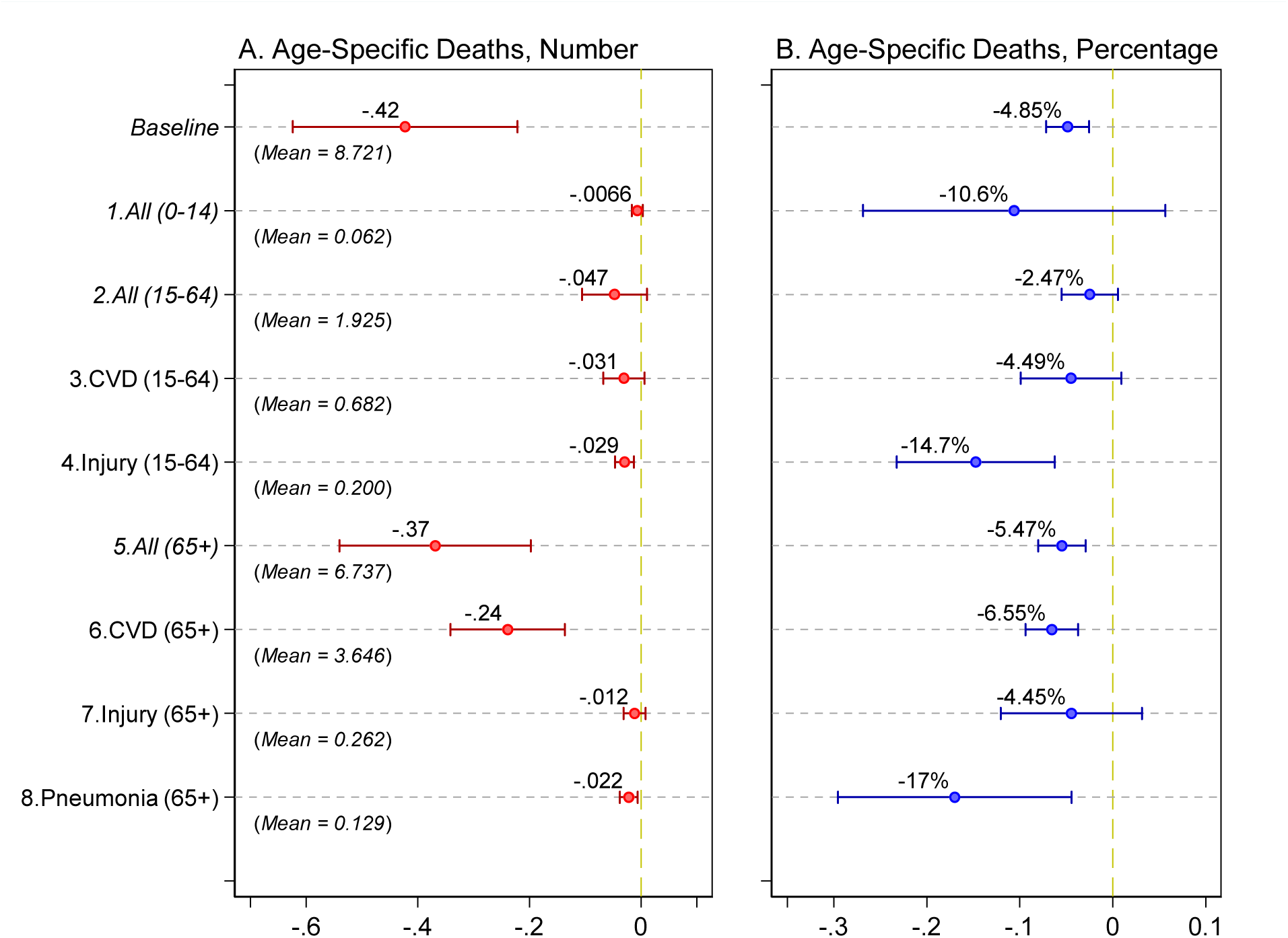
The impacts of city/community lockdowns on deaths: by age group. We examine the impacts of lockdowns on the number of deaths from different age groups. Each row represents a separate DiD regression. DSP fixed effect and date fixed effect are included in all the regressions. The number of observations for each regression is 44,548 covering 602 DSP counties except for 3 DSP sites in Wuhan. Corresponding results are reported in Table SM9.

## Supplementary Tables

**Table SM1.** Summary statistics.

|  | (1) | (2) |
| --- | --- | --- |
| **Panel A: Daily Mortality (Person)** |  |  |
| Total | 8.72 | [7.32] |
| CVD | 4.33 | [4.02] |
| Injury | 0.48 | [0.85] |
| Pneumonia | 0.15 | [0.45] |
| Neoplasms | 1.94 | [2.18] |
| Chronic respiratory diseases | 0.76 | [1.19] |
| Others | 1.07 | [1.37] |
| **Panel B: Other Variables** |  |  |
| Number of hospital bed per thousand people | 15.95 | [31.33] |
| PM<sub>2.5</sub> concentration (µg/m<sup>3</sup>) | 50.14 | [40.68] |
| GDP per capita (10,000 RMB) | 5.97 | [3.89] |
| Share of employment in manufacturing (%) | 24.20 | [12.00] |
| Daily average air pressure (hPa) | 962.96 | [100.45] |
| Daily average wind speed (m/s) | 2.67 | [1.50] |
| Daily average temperature (°C) | 4.72 | [9.16] |
| Daily average humidity (%) | 69.46 | [18.38] |
| # of DSP counties | 602 |  |

**Table SM2.** The impacts of city/community lockdowns on deaths from different causes.

|  | (1) | (2) | (3) | (4) | (5) | (6) | (7) |
| --- | --- | --- | --- | --- | --- | --- | --- |
|  | <u>Total # of Deaths</u> | <u>CVD</u> | <u>Injury</u> | <u>Non-COVID-19</u><br><u>Pneumonia</u> | <u>Neoplasms</u> | <u>Chronic Respiratory</u><br><u>Diseases</u> | <u>Other Causes</u> |
|  | Mean = | Mean = | Mean = | Mean = | Mean = | Mean = | Mean = |
|  | 8.721 | 4.330 | 0.476 | 0.150 | 1.944 | 0.756 | 1.066 |
| Lockdown | -0.429***<br>[0.102] | -0.270***<br>[0.060] | -0.044***<br>[0.014] | -0.022**<br>[0.009] | -0.037<br>[0.032] | -0.032<br>[0.022] | -0.026<br>[0.023] |
| DSP Fixed Effect | YES | YES | YES | YES | YES | YES | YES |
| Date Fixed Effect | YES | YES | YES | YES | YES | YES | YES |
| Obs. | 44548 | 44548 | 44548 | 44548 | 44548 | 44548 | 44548 |
| Adjusted R-Square | 0.788 | 0.683 | 0.315 | 0.220 | 0.563 | 0.440 | 0.390 |
| # of DSP Counties | 602 | 602 | 602 | 602 | 602 | 602 | 602 |

Each cell in the table represents a separate DiD regression. All DSP districts/counties are included in the analysis except 3 from Wuhan. The outcome variable is the daily number of non-COVID-19 deaths from the DSP districts/counties. We use mortality data from January 1 to March 14, 2020 for this analysis; the data were extracted from the DSP system on May 15, 2020. The explanatory variable is a dummy indicating whether a city associated with a DSP had a lockdown policy on a particular date. The standard errors clustered at the DSP level are reported below the estimates. \* significant at 10% \*\* significant at 5% \*\*\* significant at 1%.

**Table SM3.** Event study estimation results.

|  | (1) | (2) | (3) | (4) |
| --- | --- | --- | --- | --- |
|  | Total | CVD | Injury | Non-COVID-19<br>Pneumonia |
| >= 7 Weeks Before | 0.191<br>[0.894] | 0.476<br>[0.621] | 0.063<br>[0.117] | -0.01<br>[0.032] |
| 6 Weeks Before | -0.165<br>[0.300] | -0.159<br>[0.183] | 0.041<br>[0.040] | 0.007<br>[0.023] |
| 5 Weeks Before | -0.089<br>[0.198] | -0.112<br>[0.123] | 0.04<br>[0.029] | 0.013<br>[0.019] |
| 4 Weeks Before | -0.08<br>[0.154] | -0.09<br>[0.100] | 0.028<br>[0.025] | 0.033**<br>[0.016] |
| 3 Weeks Before | 0.049<br>[0.130] | -0.034<br>[0.089] | 0.061***<br>[0.023] | 0.029**<br>[0.013] |
| 2 Weeks Before | -0.068<br>[0.105] | -0.055<br>[0.069] | 0.036*<br>[0.021] | 0.035***<br>[0.013] |
| Week of Lockdown Policy | -0.213**<br>[0.092] | -0.169**<br>[0.065] | -0.01<br>[0.018] | -0.007<br>[0.013] |
| 1 Week Later | -0.329***<br>[0.117] | -0.272***<br>[0.079] | -0.019<br>[0.021] | 0.003<br>[0.013] |
| 2 Weeks Later | -0.524***<br>[0.134] | -0.364***<br>[0.086] | -0.029<br>[0.024] | 0.011<br>[0.014] |
| 3 Weeks Later | -0.749***<br>[0.166] | -0.491***<br>[0.102] | -0.008<br>[0.024] | -0.012<br>[0.013] |
| 4 Weeks Later | -0.906***<br>[0.186] | -0.525***<br>[0.113] | -0.041<br>[0.027] | -0.016<br>[0.014] |
| 5 Weeks Later | -1.301***<br>[0.232] | -0.689***<br>[0.141] | -0.073**<br>[0.032] | -0.033**<br>[0.017] |
| >= 6 Weeks Later | -1.314***<br>[0.361] | -0.792***<br>[0.224] | -0.016<br>[0.066] | -0.050**<br>[0.025] |
| DSP Fixed Effect | YES | YES | YES | YES |
| Date Fixed Effect | YES | YES | YES | YES |
| Obs. | 44548 | 44548 | 44548 | 44548 |
| Adjusted R-Square | 0.789 | 0.683 | 0.315 | 0.220 |
| # of DSP Counties | 602 | 602 | 602 | 602 |

Notes: We include leads and lags of the start of the lockdown dummy in the regressions to test the parallel trend assumption. The dummy variable indicating one week before the lockdown is omitted from the regressions. The standard errors clustered at the DSP level are reported below the estimates. \* significant at 10% \*\* significant at 5% \*\*\* significant at 1%.

**Table SM4.** The impacts of lockdowns on cause-specific deaths.

|  | (1) | (2) | (3) | (4) |
| --- | --- | --- | --- | --- |
| *Panel A: CVD* |  |  |  |  |
|  | <u>CVD</u> | <u>Myocardial Infarction</u> | <u>Stroke</u> | <u>Other CVDs</u> |
|  | Mean = 4.330 | Mean = 1.769 | Mean = 1.818 | Mean = 0.743 |
| Lockdown | -0.270*** | -0.128*** | -0.086*** | -0.056*** |
|  | [0.060] | [0.032] | [0.033] | [0.022] |
| *Panel B: Injury* |  |  |  |  |
|  | <u>Injury</u> | <u>Traffic</u> | <u>Suicide</u> | <u>Other Injuries</u> |
|  | Mean = 0.476 | Mean = 0.102 | Mean = 0.073 | Mean = 0.301 |
| Lockdown | -0.044*** | -0.023*** | 0.003 | -0.024** |
|  | [0.014] | [0.006] | [0.005] | [0.012] |
| *Panel C: Non-COVID-19 Pneumonia* |  |  |  |  |
|  | <u>Pneumonia</u> | <u>Pneumonia Organism</u> | <u>Viral Pneumonia &</u><br><u>Bacterial Pneumonia</u> | <u>Pulmonary Infection</u> |
|  | Mean = 0.150 | Mean = 0.065 | Mean = 0.037 | Mean = 0.061 |
| Lockdown | -0.022** | -0.023*** | 0.002 | -0.001 |
|  | [0.009] | [0.006] | [0.006] | [0.004] |

Notes: Each cell in the table represents a separate DiD regression. All DSP districts/counties are included in the analysis except 3 DSP counties/districts from Wuhan. The outcome variable is the daily number of non-COVID-19 deaths from the DSP districts/counties. We use mortality data from January 1 to March 14 2020 for this analysis; the data were extracted from the DSP system on May 15 2020. The explanatory variable is a dummy indicating whether a city associated with a DSP had a lockdown policy on a particular date. DSP fixed effect and date fixed effect are both included in each regression. The total number of observations for each regression is 44,548 covering 602 DSP counties. The standard errors clustered at the DSP level are reported below the estimates. \* significant at 10% \*\* significant at 5% \*\*\* significant at 1%.

**Table SM5.** Robustness checks and placebo test.

|  | (1)<br>Total | (2)<br>CVD | (3)<br>Injury | (4)<br>Pneumonia |
| --- | --- | --- | --- | --- |
| *Panel A: With weather controls (R1 in Supplementary Fig 5)* |  |  |  |  |
| Lockdown | -0.426***<br>(0.102) | -0.266***<br>(0.061) | -0.042***<br>(0.014) | -0.021**<br>(0.010) |
| *Panel B: With weather and socio-economic controls (R2 in Supplementary Fig 5)* |  |  |  |  |
| Lockdown | -0.391***<br>[0.101] | -0.250***<br>[0.060] | -0.043***<br>[0.014] | -0.019**<br>[0.009] |
| *Panel C: Weighting by populations (R3 in Supplementary Fig 5)* |  |  |  |  |
| Lockdown | -0.431**<br>(0.176) | -0.290***<br>(0.102) | -0.041*<br>(0.024) | -0.026<br>(0.017) |
| *Panel D: Dropping cities in Hubei (R4 in Supplementary Fig 5)* |  |  |  |  |
| Lockdown | -0.455***<br>(0.106) | -0.295***<br>(0.062) | -0.037***<br>(0.014) | -0.028***<br>(0.009) |
| *Panel E: Include Wuhan (R5 in Supplementary Fig 5)* |  |  |  |  |
| Lockdown | -0.254<br>[0.159] | -0.226***<br>[0.067] | -0.038**<br>[0.015] | 0.050<br>[0.050] |
| *Panel F: Change the definition of lockdown* |  |  |  |  |
| Only city lockdown | -0.250<br>[0.193] | 0.002<br>[0.125] | -0.125***<br>[0.043] | 0.046<br>[0.034] |
| City+community lockdown | -0.448***<br>[0.106] | -0.299***<br>[0.062] | -0.035**<br>[0.014] | -0.029***<br>[0.009] |
| *Panel G: Use 2019 deaths* |  |  |  |  |
| Lockdown | 0.088<br>[0.101] | 0.102<br>[0.063] | 0.002<br>[0.016] | -0.011<br>[0.009] |

Notes: Each cell in the table represents a separate DiD regression. In Panel A, the regression includes weather controls: daily average temperature, humidity, wind speed, and air pressure. In Panel B, the regression includes socio-economic status controls: interactions between time-invariant variables and a third-order polynomial function of time. In Panel C, the regression is weighted by the population. In Panel D, DSPs in Hubei province are excluded, while, in Panel E, three DSPs in Wuhan are included in the regression. DSP fixed effect and date fixed effect are both included in each regression. These results are presented in Figure SM5. In Panel F, we define lockdown in a different way. The DSPs are classified into 1) being placed under the city lockdown, 2) being placed under both the city lockdown and the community lockdown, or 3) without any lockdowns. DSP fixed effect and date fixed effect are both included in each regression. Panel G uses day-to-day matched 2019 death tolls as dependent variables and includes the controls of a set of time-varying variables. DSP fixed effect and date fixed effect are both included in each regression. The total number of observations for each regression is 44,548 covering 602 DSP counties. The standard errors clustered at the DSP level are reported below the estimates. \* significant at 10% \*\* significant at 5% \*\*\* significant at 1%.

**Table SM6.**
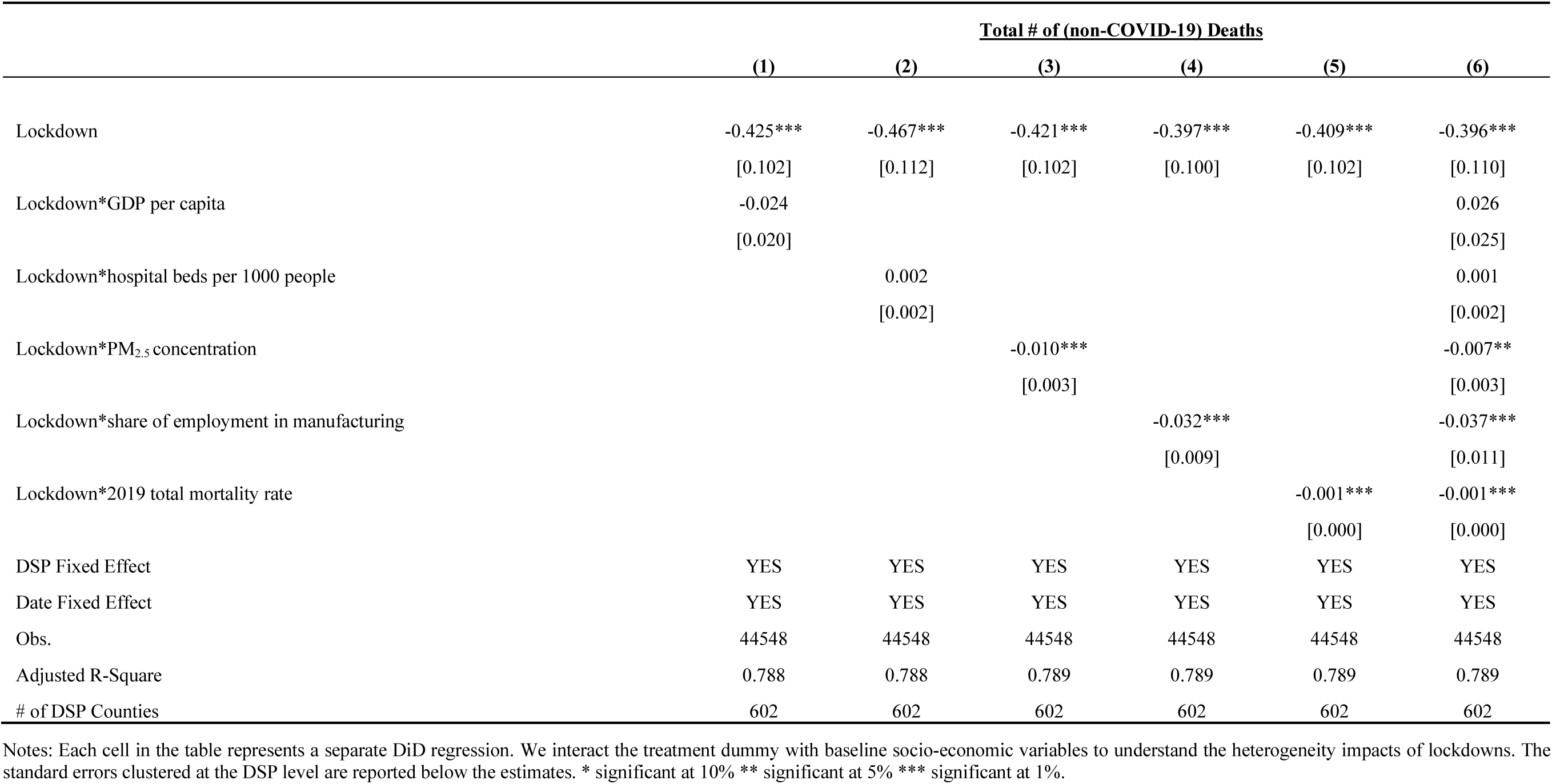
The heterogeneous impacts of city/community lockdowns on deaths.

|  | <u>Total # of (non-COVID-19) Deaths</u> |  |  |  |  |  |
| --- | --- | --- | --- | --- | --- | --- |
|  | (1) | (2) | (3) | (4) | (5) | (6) |
| Lockdown | -0.425***<br>[0.102] | -0.467***<br>[0.112] | -0.421***<br>[0.102] | -0.397***<br>[0.100] | -0.409***<br>[0.102] | -0.396***<br>[0.110] |
| Lockdown*GDP per capita | -0.024<br>[0.020] |  |  |  |  | 0.026<br>[0.025] |
| Lockdown*hospital beds per 1000 people |  | 0.002<br>[0.002] |  |  |  | 0.001<br>[0.002] |
| Lockdown*PM<sub>2.5</sub> concentration |  |  | -0.010***<br>[0.003] |  |  | -0.007**<br>[0.003] |
| Lockdown*share of employment in manufacturing |  |  |  | -0.032***<br>[0.009] |  | -0.037***<br>[0.011] |
| Lockdown*2019 total mortality rate |  |  |  |  | -0.001***<br>[0.000] | -0.001***<br>[0.000] |
| DSP Fixed Effect | YES | YES | YES | YES | YES | YES |
| Date Fixed Effect | YES | YES | YES | YES | YES | YES |
| Obs. | 44548 | 44548 | 44548 | 44548 | 44548 | 44548 |
| Adjusted R-Square | 0.788 | 0.788 | 0.789 | 0.789 | 0.789 | 0.789 |
| # of DSP Counties | 602 | 602 | 602 | 602 | 602 | 602 |

Notes: Each cell in the table represents a separate DiD regression. We interact the treatment dummy with baseline socio-economic variables to understand the heterogeneity impacts of lockdowns. The standard errors clustered at the DSP level are reported below the estimates. \* significant at 10% \*\* significant at 5% \*\*\* significant at 1%.

**Table SM7.** The heterogeneous impacts of lockdowns on CVD deaths.

|  | <u># of CVD Deaths</u> |  |  |  |  |  |
| --- | --- | --- | --- | --- | --- | --- |
|  | (1) | (2) | (3) | (4) | (5) | (6) |
| Lockdown | -0.270***<br>[0.060] | -0.311***<br>[0.065] | -0.264***<br>[0.060] | -0.258***<br>[0.060] | -0.259***<br>[0.060] | -0.276***<br>[0.064] |
| Lockdown*GDP per capita | 0.002<br>[0.010] |  |  |  |  | 0.016<br>[0.014] |
| Lockdown*hospital beds per 1000 people |  | 0.002**<br>[0.001] |  |  |  | 0.002<br>[0.001] |
| Lockdown*PM<sub>2.5</sub> concentration |  |  | -0.008***<br>[0.002] |  |  | -0.006***<br>[0.002] |
| Lockdown*share of employment in manufacturing |  |  |  | -0.012***<br>[0.004] |  | -0.015***<br>[0.005] |
| Lockdown*2019 mortality rate of CVD |  |  |  |  | -0.001***<br>[0.000] | -0.001***<br>[0.000] |
| DSP Fixed Effect | YES | YES | YES | YES | YES | YES |
| Date Fixed Effect | YES | YES | YES | YES | YES | YES |
| Obs. | 44548 | 44548 | 44548 | 44548 | 44548 | 44548 |
| # of DSP Counties | 602 | 602 | 602 | 602 | 602 | 602 |

Notes: Each cell in the table represents a separate DiD regression. We interact the treatment dummy with baseline socio-economic variables to understand the heterogeneity impacts of lockdowns. The standard errors clustered at the DSP level are reported below the estimates. \* significant at 10% \*\* significant at 5% \*\*\* significant at 1%.

**Table SM8.** The heterogeneous impacts of lockdown on deaths from injuries.

|  | <u># of Deaths from Injuries</u> |  |  |  |  |  |
| --- | --- | --- | --- | --- | --- | --- |
|  | (1) | (2) | (3) | (4) | (5) | (6) |
| Lockdown | -0.043***<br>[0.014] | -0.046***<br>[0.015] | -0.044***<br>[0.014] | -0.041***<br>[0.014] | -0.042***<br>[0.014] | -0.035**<br>[0.015] |
| Lockdown*GDP per capita | -0.004<br>[0.002] |  |  |  |  | 0.001<br>[0.003] |
| Lockdown*hospital beds per 1000 people |  | 0.000<br>[0.000] |  |  |  | -0.000<br>[0.000] |
| Lockdown*PM<sub>2.5</sub> concentration |  |  | -0.000<br>[0.000] |  |  | -0.000<br>[0.000] |
| Lockdown*share of employment in manufacturing |  |  |  | -0.003***<br>[0.001] |  | -0.003***<br>[0.001] |
| Lockdown*2019 mortality rate of injuries |  |  |  |  | -0.002***<br>[0.001] | -0.002***<br>[0.001] |
| DSP Fixed Effect | YES | YES | YES | YES | YES | YES |
| Date Fixed Effect | YES | YES | YES | YES | YES | YES |
| Obs. | 44548 | 44548 | 44548 | 44548 | 44548 | 44548 |
| # of DSP Counties | 602 | 602 | 602 | 602 | 602 | 602 |

Notes: Each cell in the table represents a separate DiD regression. We interact the treatment dummy with baseline socio-economic variables to understand the heterogeneity impacts of lockdowns. The standard errors clustered at the DSP level are reported below the estimates. \* significant at 10% \*\* significant at 5% \*\*\* significant at 1%.

**Table SM9.** The heterogeneous impacts of lockdown on deaths from pneumonia.

|  | <u># of Deaths from Non-COVID-19 Pneumonia</u> |  |  |  |  |  |
| --- | --- | --- | --- | --- | --- | --- |
|  | (1) | (2) | (3) | (4) | (5) | (6) |
| Lockdown | -0.020**<br>[0.009] | -0.004<br>[0.010] | -0.022**<br>[0.009] | -0.020**<br>[0.009] | -0.020**<br>[0.009] | -0.014<br>[0.010] |
| Lockdown*GDP per capita | -0.008***<br>[0.002] |  |  |  |  | -0.004**<br>[0.002] |
| Lockdown*hospital beds per 1000 people |  | -0.001***<br>[0.000] |  |  |  | -0.000<br>[0.000] |
| Lockdown*PM<sub>2.5</sub> concentration |  |  | 0.000<br>[0.000] |  |  | -0.000<br>[0.000] |
| Lockdown*share of employment in manufacturing |  |  |  | -0.001***<br>[0.001] |  | -0.000<br>[0.001] |
| Lockdown*2019 mortality rate of pneumonia |  |  |  |  | -0.004***<br>[0.001] | -0.004***<br>[0.001] |
| DSP Fixed Effect | YES | YES | YES | YES | YES | YES |
| Date Fixed Effect | YES | YES | YES | YES | YES | YES |
| Obs. | 44548 | 44548 | 44548 | 44548 | 44548 | 44548 |
| # of DSP Counties | 602 | 602 | 602 | 602 | 602 | 602 |

Notes: Each cell in the table represents a separate DiD regression. We interact the treatment dummy with baseline socio-economic variables to understand the heterogeneity impacts of city lockdowns. The standard errors clustered at the DSP level are reported below the estimates. \* significant at 10% \*\* significant at 5% \*\*\* significant at 1%.

**Table SM10.** The impacts of city/community lockdowns on deaths: by age group.

|  | <u>Age 0-14</u> | <u>Age 15-65</u> | <u>Age >65</u> |
| --- | --- | --- | --- |
|  | (1) | (2) | (3) |
| **Panel A. Total # of non-COVID-19 Deaths** |  |  |  |
|  | <u>Y Mean = 0.062</u> | <u>Y Mean = 1.925</u> | <u>Y Mean = 6.737</u> |
| Lockdown | -0.007 | -0.047 | -0.369*** |
|  | [0.005] | [0.030] | [0.087] |
| Adjusted R-Square | 0.041 | 0.452 | 0.767 |
| **Panel B. # of Deaths from CVD** |  |  |  |
|  | <u>Y Mean = 0.002</u> | <u>Y Mean = 0.682</u> | <u>Y Mean = 3.646</u> |
| Lockdown | -0.000 | -0.031 | -0.239*** |
|  | [0.001] | [0.019] | [0.052] |
| Adjusted R-Square | 0.005 | 0.260 | 0.663 |
| **Panel C. # of Deaths from Injuries** |  |  |  |
|  | <u>Y Mean = 0.014</u> | <u>Y Mean = 0.200</u> | <u>Y Mean = 0.262</u> |
| Lockdown | -0.003 | -0.029*** | -0.012 |
|  | [0.002] | [0.009] | [0.010] |
| Adjusted R-Square | 0.0141 | 0.124 | 0.293 |
| **Panel D. # of Deaths from Non-COVID-19 Pneumonia** |  |  |  |
|  | <u>Y Mean = 0.003</u> | <u>Y Mean = 0.018</u> | <u>Y Mean = 0.129</u> |
| Lockdown | -0.003** | 0.003 | -0.022*** |
|  | [0.001] | [0.004] | [0.008] |
| Adjusted R-Square | 0.006 | 0.024 | 0.215 |

Notes: We examine the impacts of lockdowns on the number of deaths from different age groups in this table. Each cell represents a separate DiD regression. DSP fixed effect and date fixed effect are both included in each regression. The number of observations for each regression is 44,548 covering 602 DSP counties. The standard errors clustered at the DSP level are reported below the estimates. \* significant at 10% \*\* significant at 5% \*\*\* significant at 1%.

**Table SM11.**
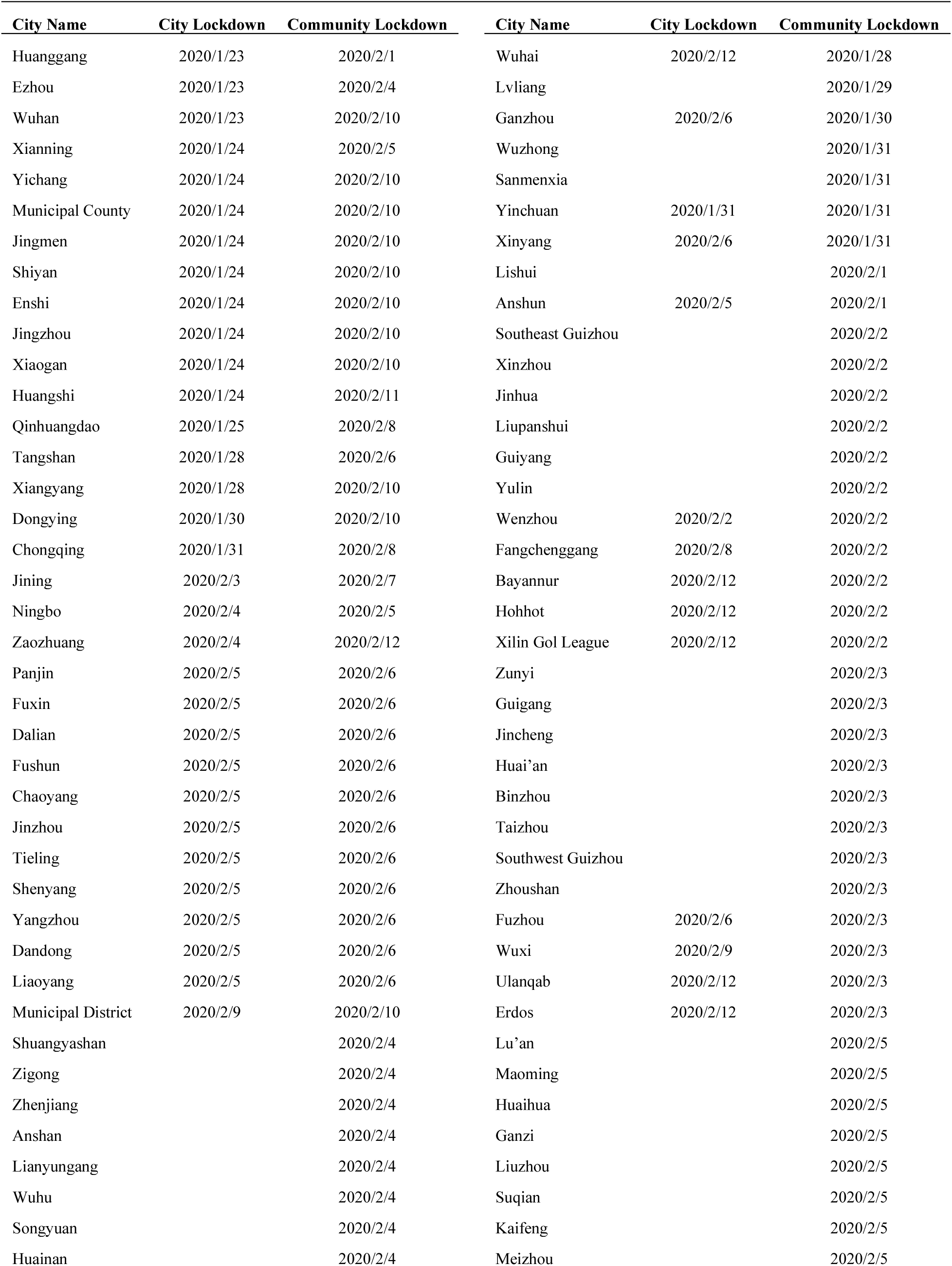
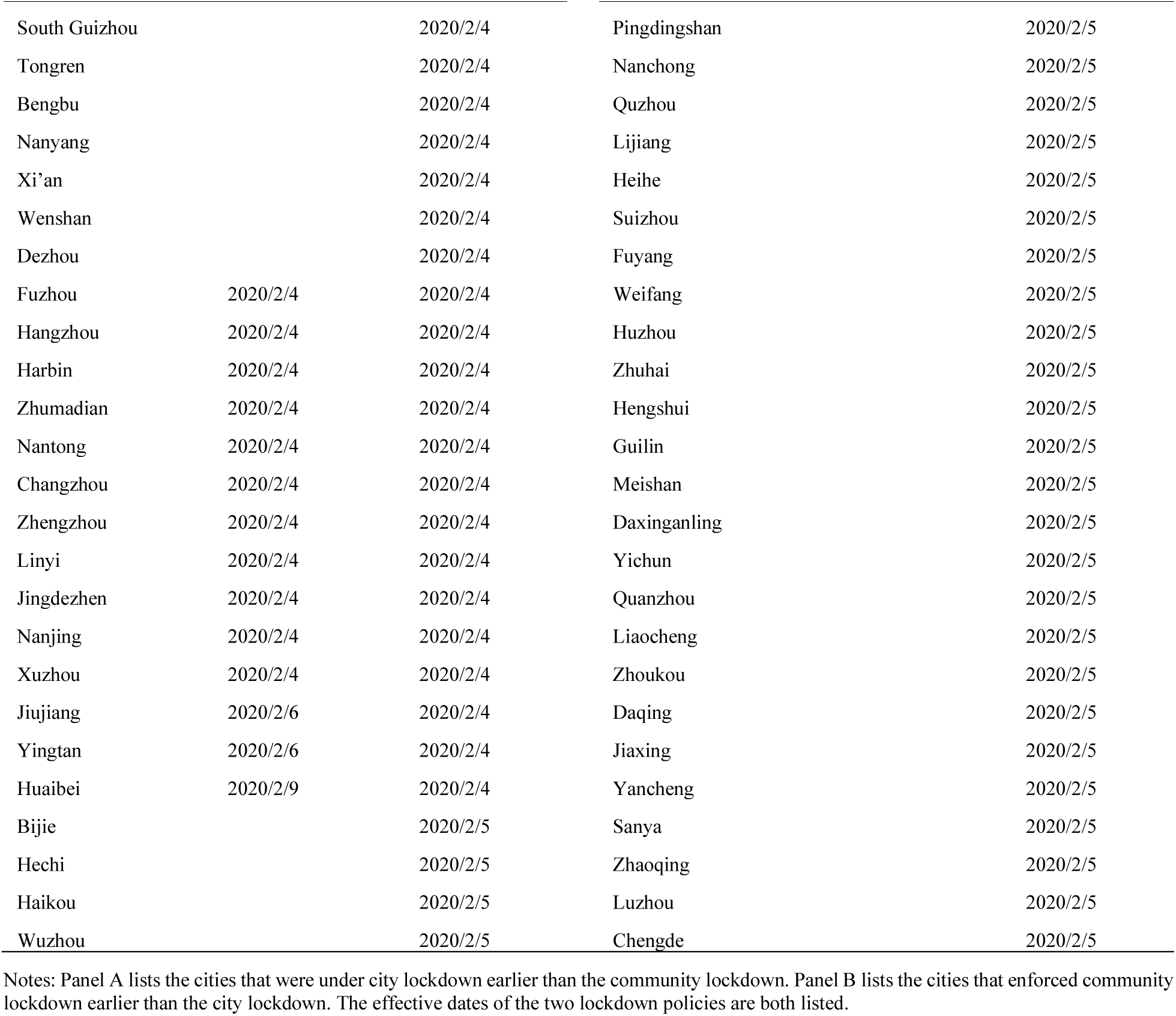
Lockdown status of Chinese cities.

## References

1. World Health Organization, WHO Coronavirus Disease (COVID-19) Dashboard. https://www.who.int/emergencies/diseases/novel-coronavirus-2019 (accessed November 2, 2020)

2. Tian H, Liu Y, Li Y, et al. An investigation of transmission control measures during the first 50 days of the COVID-19 epidemic in China. Science 2020; 368: 638–42.

3. Qiu Y, Chen X, Shi W. Impacts of social and economic factors on the transmission of coronavirus disease 2019 (COVID-19) in China. J Popul Econ 2020: 1–46.

4. Chinazzi M, Davis JT, Ajelli M, et al. The effect of travel restrictions on the spread of the 2019 novel coronavirus (COVID-19) outbreak. Science 2020; 368: 395–400.

5. Hsiang S, Allen D, Annan-Phan S, et al. The effect of large-scale anti-contagion policies on the COVID-19 pandemic [published online ahead of print, 2020 Jun 8]. Nature 2020.

6. Lai S, Ruktanonchai NW, Zhou L, et al. Effect of non-pharmaceutical interventions to contain COVID-19 in China [published online ahead of print, 2020 May 4]. Nature 2020.

7. Coibion, Gorodnichenko, Yuriy O, Webe M. Labor Markets During the Covid-19 Crisis: A preliminary view. National Bureau of Economic Research 2020.

8. Gordon SH, Sommers BD. Recessions, Poverty, and Mortality in the United States: 1993- 2012. Am J Health Econ 2016; 3: 489–510.

9. Baird S, Friedman J, Schady N. Aggregate Income Shocks and Infant Mortality in the Developing World. Rev Econ Stat 2011; 3: 847–56.

10. Jérôme A, Von Gaudecker H, Bank J. The Impact of Income Shocks on Health: Evidence from Cohort Data. J Eur Econ Assoc 2009;6: 1361–99.

11. Snyder SE, Evans WN. The effect of income on mortality: evidence from the social security notch. Rev Econ Stat 2006; 3: 482–95.

12. Cutler DM, Knaul F, Lozano R, Zurita OMAB. Financial crisis, health outcomes and ageing: Mexico in the 1980s and 1990s. J Public Econ 2002;(NO.2): 279–303.

13. Vlachadis N, Vrachnis N, Ktenas E, Vlachadi M, Kornarou E. Mortality and the economic crisis in Greece. Lancet 2014; 383: 691.

14. Falagas ME, Vouloumanou EK, Mavros MN, Karageorgopoulos DE. Economic crises and mortality: a review of the literature. Int J Clin Pract 2009; 63: 1128–35.

15. Lundin A, Lundberg I, Hallsten L, Ottosson J, Hemmingsson T. Unemployment and mortality--a longitudinal prospective study on selection and causation in 49321 Swedish middle-aged men. J Epidemiol Community Health 2010; 64: 22–8.

16. World Health Organization, COVID-19 significantly impacts health services for noncommunicable diseases. https://www.who.int/news-room/detail/01-06-2020-covid-19-significantly-impacts-health-services-for-noncommunicable-diseases(accessed July 5,2020).

17. He, G. Pan, Y. & Tanaka, T., The short-term impacts of COVID-19 lockdown on urban air pollution in China. Nature Sustainability. https://doi.org/10.1038/s41893-020-0581-y

18. Chen K, Wang M, Huang C, Kinney PL, Anastas PT. Air pollution reduction and mortality benefit during the COVID-19 outbreak in China. Lancet Planet Health. 2020;4:e210–e212.

19. Shilling F, Waetjen D. “Special Report (Update): Impact of COVID19 Mitigation on Numbers and Costs of California Traffic Crashes”. Road Ecology Center 2020 (accessed June 14, 2020).

20. Liu S, Wu X, Lopez AD, et al. An integrated national mortality surveillance system for death registration and mortality surveillance, China. Bull World Health Organ 2016; 94: 46–57.

21. Yang G, Hu J, Rao KQ, Ma J, Rao C, Lopez AD. Mortality registration and surveillance in China: History, current situation and challenges. Popul Health Metr 2005; 3: 3.

22. The Wall Street Journal, Wuhan’s Coronavirus Death Toll Surges by 50% After China Revision. https://www.wsj.com/articles/wuhans-coronavirus-death-toll-surges-by-50-after-china-reviews-data-11587110435 (accessed June 14, 2020).

23. Angrist JD, Pischke J. Mostly harmless econometrics: An empiricist’s companion. Princeton university, 2008.

24. Chen R, Yin P, Meng X, et al. Fine Particulate Air Pollution and Daily Mortality. A Nationwide Analysis in 272 Chinese Cities. Am J Respir Crit Care Med 2017; 196: 73–81.

25. Shah AS, Lee KK, McAllister DA, et al. Short term exposure to air pollution and stroke: systematic review and meta-analysis. BMJ 2015; 350: h1295.

26. Mustafic H, Jabre P, Caussin C, et al. Main air pollutants and myocardial infarction: a systematic review and meta-analysis. JAMA 2012; 307: 713–21.

27. Yin P, He G, Fan M, et al. Particulate air pollution and mortality in 38 of China’s largest cities: time series analysis. BMJ 2017; 356: j667.

28. He G, Fan M, Zhou M. The effect of air pollution on mortality in China: Evidence from the 2008 Beijing Olympic Games. Journal of Environmental Economics & Management 2016: 18–39.

29. Ebenstein A, Fan M, Greenstone M, He G, Zhou M. New evidence on the impact of sustained exposure to air pollution on life expectancy from China’s Huai River Policy. Proc Natl Acad Sci U S A 2017; 114: 10384–9.

